# Machine learning to increase the efficiency of a literature surveillance system: a performance evaluation

**DOI:** 10.1101/2023.06.18.23291567

**Authors:** Cynthia Lokker, Wael Abdelkader, Elham Bagheri, Rick Parrish, Chris Cotoi, Tamara Navarro, Federico Germini, Lori-Ann Linkins, R. Brian Haynes, Lingyang Chu, Muhammad Afzal, Alfonso Iorio

**Affiliations:** Health Information Research Unit, Department of Health Research Methods, Evidence, and Impact, McMaster University, Hamilton, Ontario, Canada; Department of Medicine, McMaster University, Hamilton, Ontario, Canada; Department of Computing and Software, McMaster University, Hamilton, Ontario, Canada; Department of Computing and Data Science, Birmingham City University, Birmingham, UK

**Keywords:** bioinformatics, machine learning, evidence-based medicine, literature retrieval, medical informatics, Natural Language Processing, biomedical informatics

## Abstract

**Background:** Given suboptimal performance of Boolean searching to identify methodologically sound and clinically relevant studies in large bibliographic databases such as MEDLINE, exploring the performance of machine learning (ML) tools is warranted.

**Objective:** Using a large internationally recognized dataset of articles tagged for methodological rigor, we trained and tested binary classification models to predict the probability of clinical research articles being of high methodologic quality to support a literature surveillance program.

**Materials and Methods:** Using an automated machine learning approach, over 12,000 models were trained on a dataset of 97,805 articles indexed in PubMed from 2012-2018 which were manually appraised for rigor by highly trained research associates with expertise in research methods and critical appraisal. As the dataset is unbalanced, with more articles that do not meet criteria for rigor, we used the unbalanced dataset and over- and under-sampled datasets. Models that maintained sensitivity for high rigor at 99% and maximized specificity were selected and tested in a retrospective set of 30,424 articles from 2020 and validated prospectively in a blinded study of 5253 articles.

**Results:** The final selected algorithm, combining a model trained in each dataset, maintained high sensitivity and achieved 57% specificity in the retrospective validation test and 53% in the prospective study. The number of articles needed to read to find one that met appraisal criteria was 3.68 (95% CI 3.52 to 3.85) in the prospective study, compared with 4.63 (95% CI 4.50 to 4.77) when relying only on Boolean searching.

**Conclusions:** ML models improved by approximately 25% the efficiency of detecting high quality clinical research publications for literature surveillance and subsequent dissemination to clinicians and other evidence users.

## INTRODUCTION

The increasing pace with which medical literature is produced is well established. So is the challenge in filtering the high-quality, clinically relevant articles from those not ready for clinical practice. Validated search strategies that filter articles by research methods, such as systematic reviews (1) and randomized controlled trials (2) have been integrated into biomedical databases to improve the efficiency of finding evidence. Though these strategies perform well, maximizing sensitivity or recall (i.e., the proportion of all on-target articles that are retrieved) comes at the cost of lower specificity (the proportion of off-target articles that are excluded from the result set) and precision (positive predictive value, i.e., the proportion of retrieved articles that are on target). Low specificity leads to significant time and resources needed to manually review and appraise the quality of the studies reported in the articles.

More recently, machine learning (ML) approaches have been applied to retrieve high quality evidence from the biomedical literature (3). There are several types of ML approaches which are determined by the mathematical method used (4,5). The most common approaches are supervised ML, unsupervised ML, ensemble learning, and neural networks. Supervised ML relies on a prelabelled training dataset to provide the machine with the necessary input to make accurate predictions (6). There are several supervised ML algorithms used to train models. For example, authors have used Artificial Neural Networks, Decision Tree, K-Nearest Neighbour, Naïve Bayes, Random Forest, and Support Vector Machine algorithms for predicting diseases (7). Automated machine learning (AutoML) iterates, selects, and optimizes ML models at multiple steps of the process (8) by automating the selection of promising algorithms, hyperparameter tuning, pre-processing, and features selection (8,9). The system searches through possible model and hyperparameter configurations and selects those that perform best on the given task. This reduces the time needed to train and test models and inaccuracies in the model that may arise from human errors and bias.

At the McMaster Health Research Information Unit (HiRU), we evaluate, at the time of publication, original studies, systematic reviews, and evidence-based guidelines in ∼120 top health care journals and research synthesis services (10) through the Premium LiteratUre Service (PLUS). Candidate studies are retrieved from PubMed daily using validated, highly sensitive Boolean search strategies to maximize recall of high-quality studies. Articles are then manually appraised by highly trained research associates with expertise in health research methods and critical appraisal to determine if they meet explicit criteria for scientific merit. Those that meet the criteria are reviewed by a clinical editor and rated for clinical relevancy and newsworthiness by a cadre of >6000 clinicians worldwide (11). The identified research is packaged into several evidence information services, tailored to the needs of knowledge users (e.g., publishers, authors, guideline developers, policy makers) and end users (e.g., clinicians) (Figure 1A). This process and application of critical appraisal criteria is consistent with the methods the HiRU team used in creating the HEDGES dataset of articles published in 2006 which has been used in the development of numerous Boolean search strategies (2,12–14) and ML models (15–17). Through PLUS, we have curated a database of articles manually classified according to methodological rigor and clinical relevance since 2012. For example, in 2019, 59 052 items indexed in PubMed in the journal set were reduced to 17 349 (29.3%) by the sensitive Boolean search filters, all of which were manually appraised by research associates. Of these, 3749 articles met critical appraisal criteria (18), giving a number of articles needed to read to identify one that met criteria (number needed to read; NNR), measured as the inverse of precision, of 4.63 (95% CI 4.50 to 4.77). The NNR provides a measure of human effort required during the critical appraisal step and a proxy for efficiency; a lower NNR reflects fewer off target articles and reduced time and effort for research associates to screen them out.

Maintaining PLUS is a resource-intensive activity, and currently limited to a subset of ∼120 journal titles (11). Reducing the NNR, by having staff focus on fewer articles that are more likely to meet criteria for rigor, while maintaining high recall (sensitivity >99%), can improve the efficiency of the process. This is particularly important as PLUS has expanded to include appraisal of all COVID-19 publications since March 2020 across all of PubMed.

Objective: To improve the efficiency of identifying high-quality clinical research to support a literature surveillance service while maintaining sensitivity at 99% (to ensure high quality articles are not missed) and reducing the NNR.

**Figure 1.**
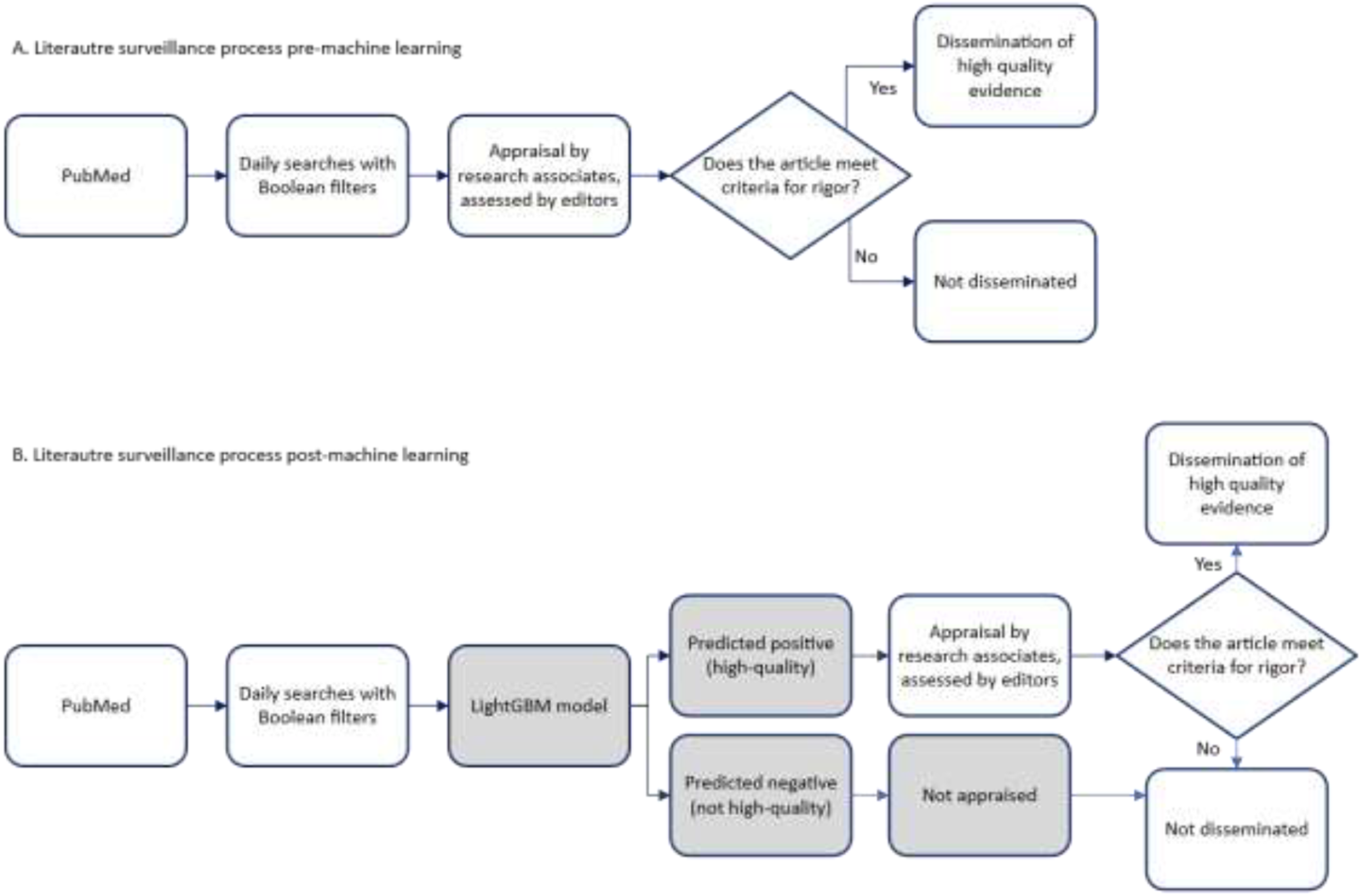
Illustration of the literature surveillance process A. before and B. after addition of a machine learning algorithm to predict quality of the article.

## MATERIALS AND METHODS

We performed a retrospective study using a labelled dataset of articles that were critically appraised for methodologic rigor and clinical relevance to train, validate, and test algorithms that predict the likelihood of a clinical article meeting appraisal criteria for rigor. We used automated ML as an efficient approach to training multiple models. Selected models were prospectively evaluated by having trained research associates, blinded to model predictions, appraise incoming articles in the literature surveillance program, as a test of the external validity of model predictions.

### Quality standard database

We define high-quality or rigor as meeting at least all critical appraisal criteria for a particular article type (review, guideline, original study) or purpose category (treatment, diagnosis, prognosis, etiology for harm primary prevention, quality improvement, economics, or clinical prediction guides) based on established evidence assessment criteria (18). The critical appraisal step, conducted manually by research associates, has previously documented high inter-rater agreement (kappa > 0.80 for all categories)(10). Articles that meet methodological criteria are then reviewed by a clinical editor with advanced research methods training and at least three members of an online community of >4000 clinicians who rate the methodologically rigorous articles for clinical relevance and newsworthiness (11). Over the course of two decades, we have reviewed more than 500,000 articles and have curated an internal database that also includes articles that did not meet methodological rigor criteria or clinical relevance or newsworthiness. Notably, the database is unbalanced, with about 4.5 times the number of articles that fail to meet methodologic rigor or clinical relevance than those that pass. The growing database now includes articles on COVID-19 indexed in PubMed not limited to the core journal set.

### Model training and performance

Our approach to model training was to use automated machine learning (AutoML), a process that allows for running multiple sequential experiments with varying settings. The process, depicted in Figure 2, automatically iterates model training using the combinations of pre-processing options, weighting methods, feature selection, and hyper-parameters listed in Table 1, and optimizes selections to identify the best performing combinations—essentially the approach optimizes performance and abandons steps that do not lead to better performing models. The performance of an AutoML system depends on the quality of the data and the specific task at hand. We chose AutoML for this study since our dataset was of high quality as it was reviewed and appraised by human experts, and we wanted to remove our biases and grow our understanding of the best approaches for our dataset. AutoML allowed for experimentation while developing expertise. We used Microsoft’s ML.NET AutoML (19) to train and test binary classification models that predicted if an article was of high-quality or not to help us get a highly optimized model, driven by a set goal of improving specificity while maintaining sensitivity above 99%.

We tested weighting by term frequency (TF), inverse document frequency (IDF), and TF-IDF to account for frequency of words within titles and abstracts of articles and their frequency across a dataset. A convenience sample of algorithms available in the public domain and in ML.NET that provided a probability score as an output measure was selected for training. This allowed us to set a threshold of 99% sensitivity rather than the default 50%. The available algorithms at the time of training were FastTree, Limited-memory Broyden-Fletcher-Goldfarb-Shanno Logistic Regression, Stochastic Dual Coordinate Ascent Logistic Regression, Stochastic Gradient Descent Calibrated Logistic Regression, Symbolic SGD Logistic Regression, and Light Gradient Boosting Machine (LightGBM) .

**Table 1.**
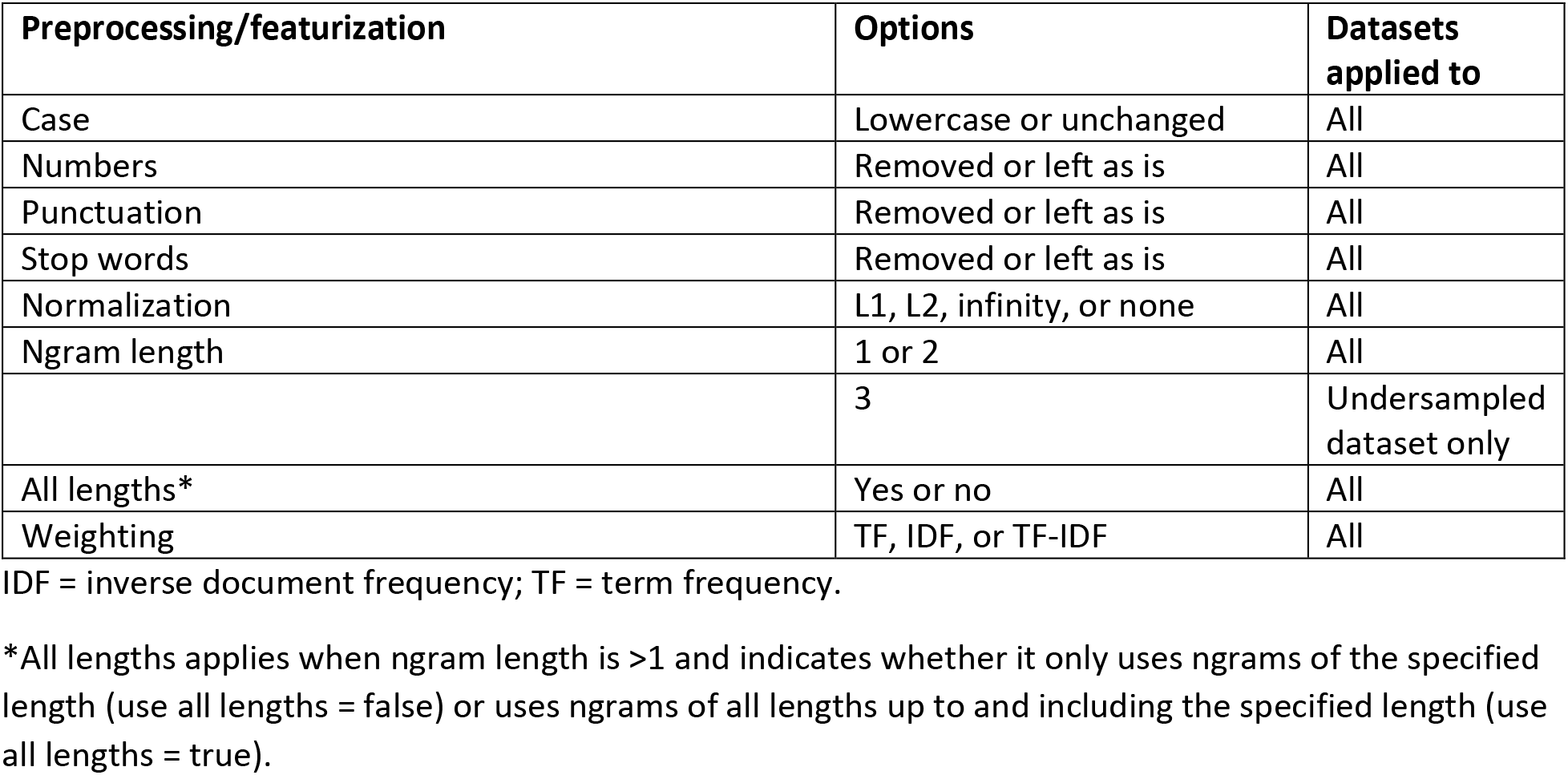
Parameters and features used in the training of models using automated ML.

| Preprocessing/featurization | Options | Datasets applied to |
| --- | --- | --- |
| Case | Lowercase or unchanged | All |
| Numbers | Removed or left as is | All |
| Punctuation | Removed or left as is | All |
| Stop words | Removed or left as is | All |
| Normalization | L1, L2, infinity, or none | All |
| Ngram length | 1 or 2 | All |
|  | 3 | Undersampled dataset only |
| All lengths* | Yes or no | All |
| Weighting | TF, IDF, or TF-IDF | All |

IDF = inverse document frequency; TF = term frequency.
\*All lengths applies when ngram length is >1 and indicates whether it only uses ngrams of the specified length (use all lengths = false) or uses ngrams of all lengths up to and including the specified length (use all lengths = true).

**Figure 2.**
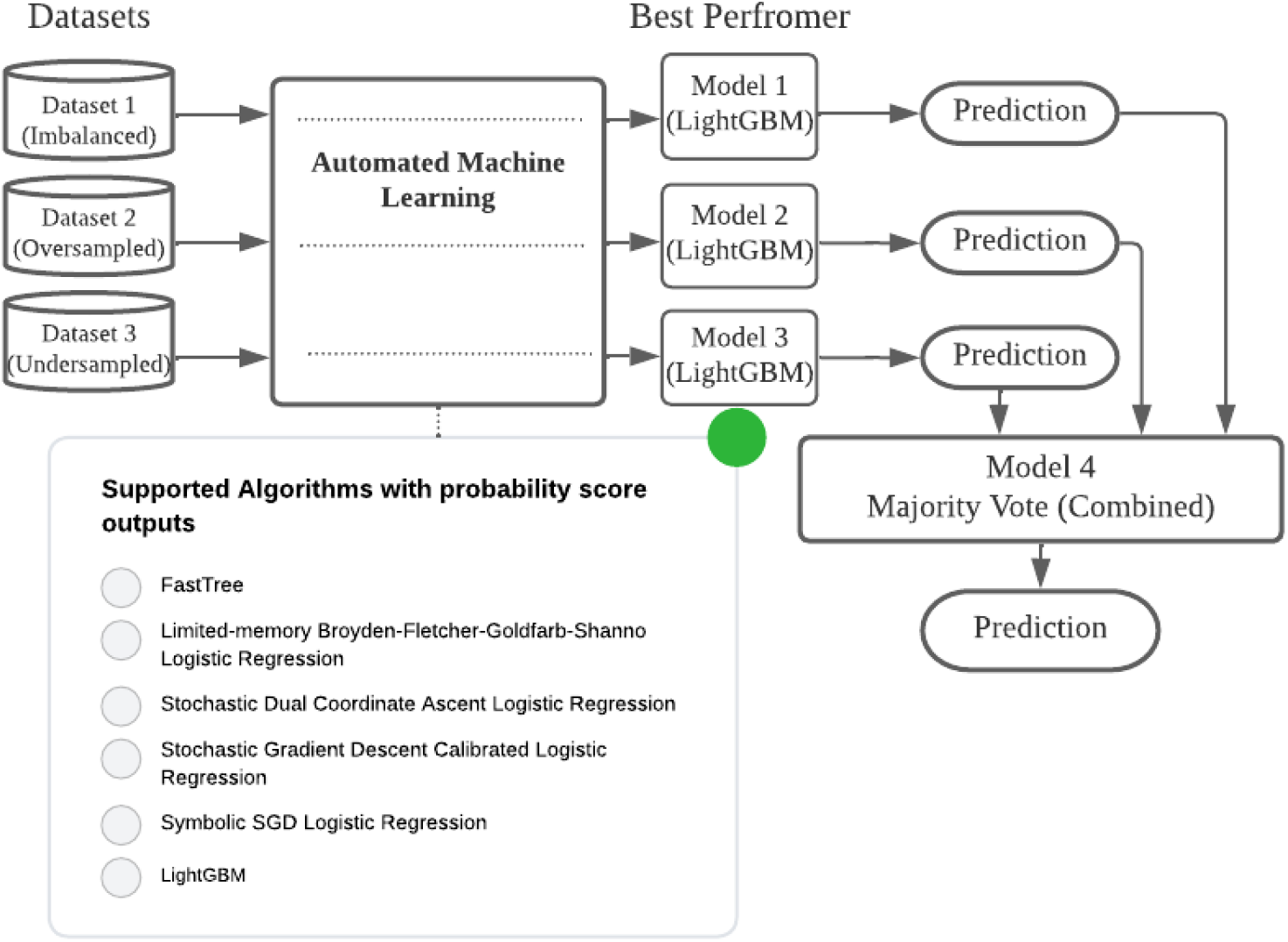
Example depiction of the autoML process.

Models with >99% sensitivity were ranked by maximal specificity. The classification models were trained using titles and abstracts of a random 80% of articles from 2012-2018 (n = 97,805). Of these, 17,824 met criteria for rigor for one or more article categories; 79,981 did not. To address the imbalance in articles, we created 3 training datasets: 80% of the full dataset (unbalanced; n = 97,805), and two additional datasets to achieve balance through oversampling (articles meeting criteria were included multiple times to equal the number of articles that did not; n = 159,962) and undersampling (random subset of articles not meeting criteria were matched to the number that did; n = 35,648).

Trained models were tested on the remaining hold-out set of 20% (n = 24,678) of articles from 2012-2018. Models with ≥99% sensitivity with the best specificity for each of the full, over-, and under-sampled datasets were retained, and one model per dataset was selected from the leaderboard. Models return a probability score ranging from 0 (does not meet criteria) to 1 (meets criteria) for each article. The probability threshold was determined as the point where sensitivity was 99%. To determine if ensembling the three models improved performance compared with the individual models, we tested their performance individually and combined—using a majority vote such that articles predicted to pass in ≥2 of the 3 models were classified as ‘pass’ (or classified as ‘fail’ if 0 or 1 model predicted a pass)—in a retrospective sample of 30,424 articles in our dataset that were published in 2020.

The performance of the models in the hold-out test set is akin to internal validation. Since our goal is to implement an algorithm into a literature surveillance program, we assessed its performance in real-time in an external test on unseen data. We prospectively evaluated the accuracy of the majority vote algorithm by applying it after Boolean searches of PubMed and before critical appraisal by our research associates, who were blinded to the predictions of 5253 articles published between March 9 to May 11, 2021. Staff appraised all articles predicted to pass and a random subset of those predicted to fail. False negative articles were assessed by a senior clinical researcher (BH) to determine clinical relevance and newsworthiness .

### Evaluation metrics

For all trained models, during the testing phase we calculated sensitivity (recall), specificity, accuracy, precision, NNR (1/precision), and F-score (harmonic mean of recall and precision metrics) in the 20% hold-out set of articles from 2012-2018. We also calculated the area-under-the-curve (AUC) of the receiver operating characteristic (ROC) curve. The ROC curve is created by plotting the true positive rate (sensitivity) against the false positive rate (1-specificity) by varying the threshold applied to the probability outputs of a classifier. AUC is thus a threshold-independent parameter which demonstrates the overall performance of the classifier. The statistical probability was calculated for the three selected models and majority vote algorithm in the 2020 data and the prospective evaluation. For the prospective evaluation, we estimated the bias-corrected sensitivity and specificity with corresponding 95% confidence intervals (CIs) using the Begg and Greenes (20) formula that corrects for any bias when a only subsample is verified to account for the articles that were predicted to fail and that were not verified by design. The bias correction models the diagnostic distribution of the articles that were verified (20).

## RESULTS

### Selected models and their performance

We trained 3456 models using the unbalanced and oversampled datasets and 5760 models using the undersampled dataset. The preprocessing steps and parameters used in the selected top performing models are shown in Table 2; each of the three selected models used the LightGBM binary classification algorithm (21,22). LightGBM (light gradient-boosting machine)(21) is a gradient boosting framework that uses decision tree algorithms. It is a more efficient implementation of gradient boosting decision tree (23) which is an ensemble model of decision trees trained in sequence and a widely-used machine learning algorithm, thanks to its efficiency, accuracy, and interpretability. The performance characteristics of each of the three models in the test datasets from 2012-18 and 2020 are listed in Table 3. The oversampled dataset shows more variation in the ROC curves of all trained classifiers, which could be due to having higher number of training examples resulting in underfitting of some classifiers; the ROC curves are available in Appendix A. The classifiers trained on undersampled data also have slightly more variation in performance compared to unbalanced data, which may be because some information was lost compared to using all available data. Nevertheless, the AUC values for the three top performing models are very close to each other indicating a high performance for the selected LightGBM model in all three cases.

**Table 2.** Characteristics of the dataset, preprocessing, and feature extraction steps employed by AutoML in the training of the model selected from each dataset experiment*

|  | **Model 1**<br>**(Unbalanced**<br>**dataset)** | **Model 2**<br>**(Balanced by**<br>**over-sampling)** | **Model 3**<br>**(Balanced by**<br>**under-**<br>**sampling)** |
| --- | --- | --- | --- |
| Number of articles in training datasets | 97 805 | 159 962 | 35 648 |
| Ratio of negative:positive articles (or %positive) | 4.5:1 | 1:1 | 1:1 |
| Number of models trained | 3456 | 3456 | 5760 |
| **Features employed in the selected best model:** |  |  |  |
| Text converted to lowercase | Yes | Yes | No |
| Removal of punctuation | Yes | Yes | Yes |
| Removal of stop words | Yes | No | No |
| Removal of Diacritics | Yes | Yes | Yes |
| Removal of numbers | Yes | Yes | Yes |
| Weighting method | TF-IDF | TF-IDF | TF-IDF |
| Normalization technique | None | None | L1 |
| N-grams | Uni-grams | Bi-grams | Tri-grams |

L1 = Manhattan Distance or Taxicab norm. TF-IDF = term frequency - inverse document frequency.
\*All selected models used LightGBM binary classification model.

**Table 3.** Performance characteristics for the three models in the testing datasets (20% from 2012-2018, and 2020).

| Testing dataset | Model | Sensitivity (95% CI) | Specificity (CI) | Precision | F-score | Accuracy | NNR (CI) | AUC (CI) |
| --- | --- | --- | --- | --- | --- | --- | --- | --- |
| 2012-2018* | 2 | 99.0% (98.7 to 99.3) | 53.7% (53.0 to 54.4) | 32.6% | 0.490 | 62.1% | 3.07 (3.02 to 3.13) | 0.952 (0.949 to 0.956) |
|  | 1 | 99.0% (98.7 to 99.3) | 51.0% (50.3 to 51.6) | 31.3% | 0.475 | 59.8% | 3.20 (3.14 to 3.26) | 0.952 (0.949 to 0.955) |
|  | 3 | 99.0% (98.7 to 99.3) | 51.8% (51.1 to 52.5) | 31.6% | 0.480 | 60.5% | 3.16 (3.10 to 3.22) | 0.948 (0.944 to 0.951) |
| 2020† | Combined‡ | 99.2% (98.8 to 99.4) | 57.5% (56.9 to 58.1) | 26.2% | 0.415 | 63.0% | 3.86 (3.79 to 3.93) | NA |
|  | 2 | 99.1% (98.7 to 99.4) | 57.3% (56.7 to 57.9) | 26.1% | 0.413 | 62.8% | 3.87 (3.80 to 3.95) | 0.962 (0.959 to 0.964) |
|  | 1 | 99.0% (98.7 to 99.3) | 56.4% (55.8 to 57.0) | 25.7% | 0.408 | 62.0% | 3.94 (3.86 to 4.01) | 0.959 (0.956 to 0.962) |
|  | 3 | 99.0% (98.7 to 99.3) | 56.0% (55.4 to 56.6) | 25.5% | 0.406 | 61.7% | 3.96 (3.88 to 4.04) | 0.956 (0.953 to 0.959) |

NA = not applicable; NNR = number needed to read. \*20% of the articles from 2012-2018 for internal testing, n=24,677. †n=30,424. ‡Predictions determined by majority vote of articles meeting 2 of 3 probability thresholds from the unbalanced, over- and under-sampled models to pass.

### Prospective evaluation

For the prospective evaluation, we opted to use the majority vote algorithm to classify 5253 consecutive articles entering the surveillance system; 2856 (54%) were predicted to be high quality and 2397 (46%) were not (Figure 3). All the 2856 predicted to be high quality and a random sample of 584 of the 2397 predicted to not be high quality were assessed by human appraisers. The remaining 1813 (90%) were not assessed and considered true negatives. Of the random sample predicted to not be high quality and appraised by staff, four were adjudicated to be high quality (false negatives), all of which required using information from the full text of the manuscript to confirm they met the appraisal criteria for their article categories. Sensitivity was 99.5% (CI, 98.7 to 99.9), specificity was 53.5% (CI, 52.0 to 55.0), and the F-score was 0.427 (Table 4). The results of the corrected analysis that adjusts for 1813 articles that were not assessed (bias corrected calculation) overlapped with the uncorrected values (Table 4).

**Figure 3.**
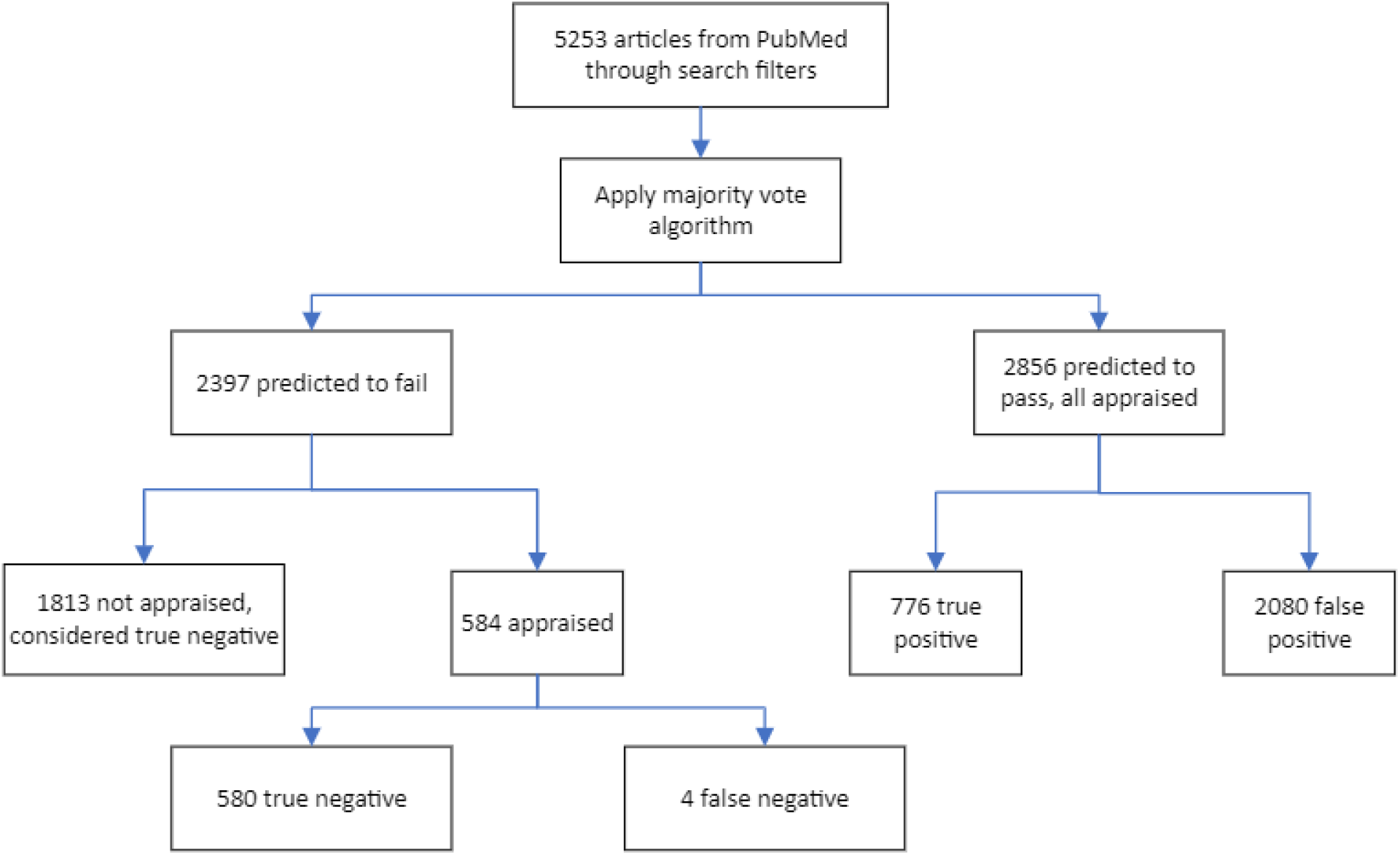
Prospective evaluation of model performance in >5000 articles retrieved from PubMed.

**Table 4.**
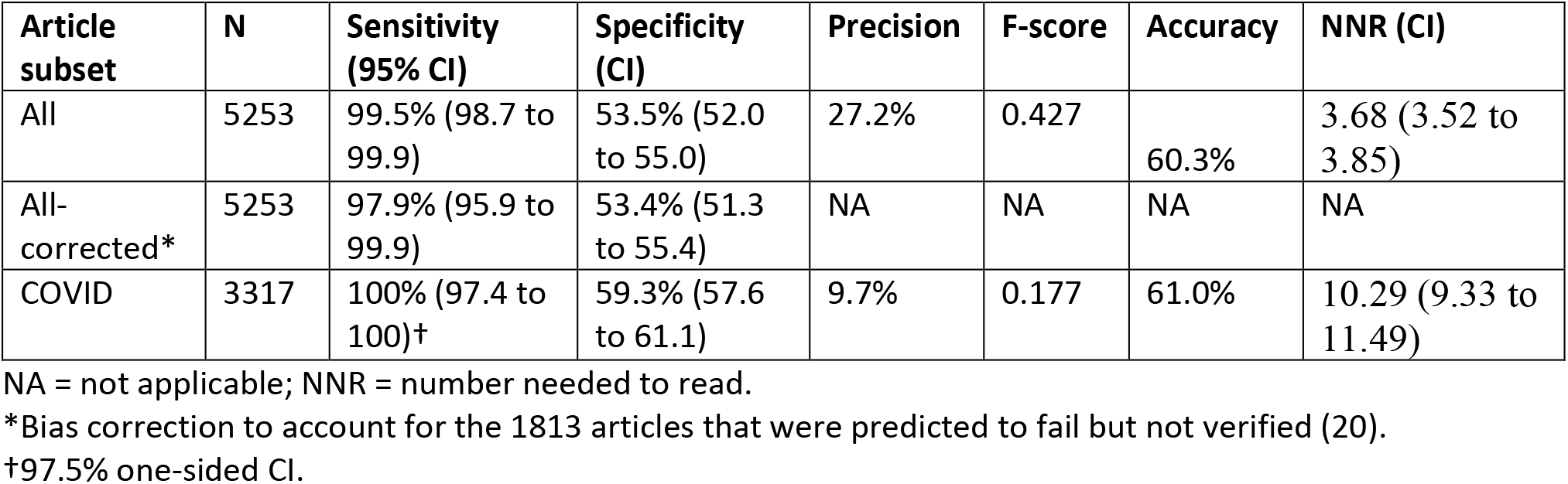
Prospective performance of the majority vote ML algorithm.

## DISCUSSION

### Model training and performance

The initial approach of using AutoML and supervised machine learning led to efficient development of models for identifying articles pre-filtered by highly sensitive Boolean searches likely to be found rigorous and clinically relevant at critical appraisal. Adopting AutoML was time efficient and allowed for the system to test various permutations of preprocessing steps and algorithms with minimal programmer time. Each of the selected highest performing models used the LightGBM binary classification algorithm (21). Our selected top performing models used TF-IDF, which accounts for both the number of times a word appears in a document and the inverse of the number of documents in the dataset that includes the word; this essentially eliminates naturally occurring English terms and gives higher values to words that are less common across the documents, or articles, in this case.

Training the models with datasets of varying size and balanced/unbalanced allowed us to assess the value in data augmentation. We also explored the effect of combining models to determine if such an approach would improve performance. Though the improvement was very small, our decision to test the ensemble and implement it was based solely on our efforts to maximize specificity to reduce the NNR. Keeping sensitivity high at 99%, the specificity of the trained models was >50% in the random test set from 2012-2018, with slightly better performance with the model trained using the larger oversampled dataset compared with the unbalanced and undersampled datasets. Though this offered a larger sample, it came at the cost of time required for model training. Despite having more models trained using the undersampled dataset, the performance of the top models was consistent with the unbalanced dataset model. All models had similar specificity in the 2020 dataset and performed marginally better than in the 2012-18 set. This could be the result of a larger sample and a broader range of journal titles and article types with the inclusion of COVID-19 publications.

The results for the majority vote combined models, where articles predicted to pass for at least two of the three models, did not factually improve the performance in the three testing datasets across years. Such ensemble approaches of combining models have been used by Aphinyanaphongs et al., (24) and Kilicoglu et al. (17) and showed improved F-scores. Ensemble techniques are used to reduce variability across models by averaging out the errors made by each, assuming they are making different errors (25). Ensemble models generally perform better when the base models they combine are as diverse as possible (26). Our three models were built to represent the full unbalanced dataset, a balanced undersampled dataset, and a larger oversampled dataset, but they include the same positive class of articles and employed the same type of ML model and are likely not diverse enough to boost performance when combined.

Testing and application of the ML models improved specificity compared with our traditional approach of Boolean filters alone. Our goal was to maximize recall/sensitivity and specificity and reduce the NNR. Prior to applying the ML models to the PLUS process (and before COVID-19), our NNR in 2019 was 4.63 (95% CI 4.50 to 4.77). With the addition of COVID-19 articles, in 2020 our overall NNR was 7.11 (CI 6.92 to 7.31). In the 2021 prospective evaluation with the addition of the ML models, the NNR was reduced to 3.68 (CI 3.52 to 3.85) for all article categories. For the four false negative articles, the main apparent reason for being missed was insufficient information in the title or abstract to be judged as valid.

### Machine learning for biomedical evidence

Our approach is consistent with reported methods in our recent systematic review of ML applied to improve the identification of high-quality articles (3). We used an established gold standard for high quality articles produced through our PLUS process. Seven studies included in the review trained their models using the Hedges dataset or articles included in ACP Journal Club, both of which are produced by the same process in HiRU (3). Like other studies, we used title and abstracts as training features. Of the 10 studies included in our earlier systematic review, seven used datasets of articles that had been critically appraised by our process (3).

Our models optimized recall to reduce the loss of relevant articles but that came at the cost of reducing specificity and precision. The precision of our models, which ranged from 26% to 33%, was surpassed by Kilicoglu et al. (17) who used ensemble models (74%), and Del Fiol et al.(16) (34%) and Afzal et al. (27) (86%) who used neural network models. The high precision achieved is likely attributed to the targeting particular categories of articles. Kilicoglu et al. (17) used an ensemble model which achieved a precision of 37% and recall of 63% when applied to articles in general, and precision of 74% and recall of 86% when used to identify rigorous treatment articles—all of which are randomized controlled trials (RCTs)— a category with established terminology and structure for reporting. Afzal et al. (27) used the Cochrane library as training dataset for their neural network, which includes systematic reviews and RCTs, which again use explicit study design terminologies in the title, abstract, or commonly both. This facilitates the retrieval function for the model and improves the overall model performance. The use of additional features, like MeSH terms and MEDLINE metadata could also explain the improved performance of their model, though these elements are not readily available for an article when it is first posted in PubMed; there is a delay from PubMed creation date and indexing being applied, a delay that varies by journal title (28). Aphinyanaphongs et al. trained models using treatment, diagnosis, prognosis, and etiology articles from ACP Journal Club (29,30) (24,31)which reflects the range of article types included in our dataset.

Deep learning is also being applied to address information retrieval and evidence classification. Ambalavanan and Devarakonda (32) trained sciBERT, a pretrained deep learning algorithm, and looked at both class ratios and size of the training sets for classifiers of treatment articles using the Clinical Hedges dataset. They found that recall was maximized when there were more positive to negative articles, precision was improved in larger training sets though there appeared to be a point at which bigger did not mean better, and the F-score was optimal using a reasonably large set of balanced articles (15,000:15,000). They modeled a number of steps in the article classification process (e.g., of interest to humans, original study, treatment article, rigorous), and found that the F-score was lowest for predicting rigor, which is a more difficult task. Notably, their study focused on articles in the treatment category while our model covers articles from the full range of categories covered in the surveillance process.

We recently published results on models for classifying articles for rigor that we developed by finetuning BioBERT, another pretrained language model (33). Our selected model outperformed the model reported here, saving 60% of the manual assessments required by research associates. That model has been integrated into our process but does require more computational power. Future work will continue with both deep and shallow learning as each has optimal uses depending on resources required for implementation.

F-score is the balance between recall (not missing a significant number of instances) and precision (how many instances it classifies correctly) and it provides an intuitive value of the robustness of the developed models. The article classification tasks assigned to the model were binary, with recall optimized to increase the model robustness over its precision. This intentional optimization towards higher recall was guided by our motive to minimize the chance of losing relevant articles. This limited our flexibility in maximizing precision and resulted in a lower overall F-score. The wide range of article categories in both training dataset and the stream of articles screened by the model would also have reduced the F-scores. Had we sought to classify articles from a particular purpose category, such as treatment studies using RCT designs, we expect the F-score would be higher.

### Implications for evidence surveillance

Retrieving the best quality evidence to clinicians has driven research into the creation of initial Boolean search strategies and now the advancements made applying ML models. We implemented the majority vote ML algorithm into our process in May 2021 (see Figure 1B). Between May 11, 2021 to Mar 11, 2022, 25 867 articles were retrieved from PubMed with the Boolean searches; 11 776 (45.5%) were predicted not to meet criteria and were removed from the critical appraisal queue. With a conservative estimated time of 5 minutes of human resources to appraise each article, this saved >981 hours of research associate time during that period while maintaining the integrity of the evidence processed.

This has been particularly important as we added COVID-19 related articles from all indexed journals to our surveillance program in 2020 to support quick access for practitioners, policy makers, and lay persons to appraised emerging research through the COVID-19 Evidence Alerts website (34). The ML model has offset some of the additional burden of this growing body of COVID-19 literature.

### Future model development

Using Auto-ML, we were able to train and test models that improved the efficiency of our literature surveillance process. There are several pretrained deep learning language models, such as BERT, BioBERT, and PubMedBERT, available for application to clinical literature. We have begun preliminary development of deep learning models using our dataset and the results are promising. Our future research includes assessing model performance by category of articles and applying our models more broadly beyond the titles monitored for PLUS. Given the richness of our dataset, including tagged reasons for not meeting critical appraisal criteria and other article metadata captured at the time of appraisal, we hope to enhance model performance by leveraging these data.

### Strengths and limitations

Our models were trained using the largest tagged dataset of health care research articles across a range of article categories to date and based on an established gold standard in the field. Although the critical appraisal criteria are applied by a single reader, all included studies and those passing with questions are assessed by a final editor. The dataset overcomes some of the challenges we identified in our review: 1) the criteria applied to assess rigor is an established gold standard based on best evidence-based medicine practices; 2) the dataset is the largest, yet, and the training dataset included 17 824 articles in the high-quality class that allowed for creating of oversampled and undersampled datasets for training; 3) journals for a range of clinical domains are included in the dataset (current list of journals can be found here: https://hiru.mcmaster.ca/hiru/journalslist.asp); and 4) the training dataset contemporary and includes articles from 2012-2018 and was tested in 2020 dataset. The prospective, blinded evaluation of the performance of the selected combined models highlights the value of real-world application and impact.

The models, however, were derived using prefiltered articles from PubMed for a subset of ∼120 journals and generalizability to all the content in a literature database is uncertain. These concerns are allayed by the performance of the models in the 2020 articles which are more numerous and cover a greater array of journal titles as all pre-filtered COVID-19 articles were included. Though the number to read was higher (not surprising given the amount of lower quality evidence in COVID-related studies), specificity and accuracy were improved. We used logistic regression approaches, and more advanced deep learning techniques expected to perform better, as seen in the results of Del Fiol et al (16) and Afzal et al (27). We have started using deep learning approaches in furthering our work in the area, initially to further increase the specificity of classifiers for all categories of articles. We plan to evaluate models for applications other than literature surveillance and investigate questions about optimal class ratios and training dataset size for model development. Further work will include training deep learning models for specific article categories, using more of the features in our dataset that correspond to rigor, and developing interpretable AI models.

### Conclusion

Using ML-based probability ranking, we improved the specificity of identifying biomedical articles that meet methodological rigor criteria while preserving a very high sensitivity. The selected models perform well in an active surveillance program that supports knowledge translation to practicing clinicians. Future work includes training deep learning models using the dataset to develop higher performing models to facilitate identification of high-quality research soon after publication.

## Data Availability

Data sharing is possible via collaboration agreements between the authors and those requesting access. The HEDGES article database is proprietary, owned by Dr Haynes via McMaster University, and can be made available with academic collaboration agreements or commercial contracts.

## Supporting Information

Appendix A. Receiver operating characteristic (ROC) curves for the models trained on the 3 datasets

